# Androgen-Related Associations With CBC-Derived Inflammatory Indices in Young Women With Polycystic Ovary Syndrome

**DOI:** 10.64898/2025.12.23.25342888

**Authors:** Natalia Piórkowska, Anna Bizoń, Kacper Bizoń, Grzegorz Franik

## Abstract

**Context:** Polycystic ovary syndrome (PCOS) is characterized by hyperandrogenism and frequent metabolic disturbances, including insulin resistance (IR), and is commonly accompanied by chronic low-grade inflammation. Complete blood count (CBC)–derived indices provide inexpensive markers reflecting systemic inflammatory and hematologic status; however, their relationships with androgen-related features in PCOS remain incompletely characterized.

**Objective:** To evaluate associations between androgen-related biomarkers and CBC-derived indices in young women with PCOS and to determine whether these associations persist after accounting for insulin resistance.

**Design:** Cross-sectional observational study.

**Setting:** Single-center Gynecological Endocrinology Clinic in Katowice, Poland.

**Participants:** Women aged 16–25 years diagnosed with PCOS (Rotterdam criteria) between 2018 and 2025 (N = 1,300). Available-case and complete-case analyses were performed.

**Main Outcome Measures:** Neutrophil-to-lymphocyte ratio (NLR), red cell distribution width (RDW), and platelet-to-lymphocyte ratio (PLR). Associations with free androgen index (FAI), total testosterone, and dehydroepiandrosterone sulfate (DHEAS) were assessed using Spearman correlation and multivariable linear regression models adjusted for age and log-transformed HOMA-IR with heteroskedasticity-consistent (HC3) standard errors. False discovery rate (FDR) correction was applied.

**Results:** FAI was positively associated with NLR in both available-case and complete-case analyses (Spearman ρ = 0.201; FDR-adjusted q < 0.001). DHEAS showed a positive association with NLR in complete-case analysis (q = 0.040).

In multivariable models adjusted for age and log(HOMA-IR) (n = 885–888 depending on outcome), higher FAI was independently associated with lower RDW (β = −0.075; 95% CI −0.141 to −0.009; q = 0.032) and lower PLR (β = −2.37; 95% CI −4.60 to −0.14; q = 0.042). Higher DHEAS was independently associated with lower RDW (β = −0.00056; 95% CI −0.00109 to −0.00004; q = 0.042). In complete-case analysis, total testosterone was inversely associated with PLR (β = −3.91; 95% CI −7.58 to −0.24; q = 0.038). Associations between androgen markers and NLR were attenuated after adjustment.

**Conclusions:** Among young women with PCOS, androgen-related biomarkers are independently associated with selected CBC-derived indices, particularly RDW and PLR, whereas associations with NLR appear partly explained by shared metabolic correlates. These findings suggest that androgen excess may be linked to subtle hematologic alterations in early-stage PCOS beyond insulin resistance.

## INTRODUCTION

Polycystic ovary syndrome (PCOS) is among the most prevalent endocrine disorders affecting women of reproductive age and is defined by hyperandrogenism, ovulatory dysfunction, and polycystic ovarian morphology [1]. Beyond its reproductive manifestations, PCOS is increasingly recognized as a multisystem disorder with metabolic and immunologic features [2]. Insulin resistance (IR) is highly prevalent and contributes substantially to long-term cardiometabolic risk [3]; however, androgen excess remains the central pathophysiologic hallmark of the syndrome and may exert systemic effects extending beyond metabolic pathways [4].

Chronic low-grade inflammation has been consistently reported in PCOS, including alterations in circulating cytokines, leukocyte subsets, and markers of innate immune activation [5–7]. Although IR is frequently implicated as a principal driver of inflammatory changes [8], experimental and translational evidence suggests that androgens themselves may modulate immune and hematopoietic processes. Androgen receptors are expressed in multiple immune cell populations and hematopoietic progenitors [9,10], and androgen signaling has been shown to influence leukocyte differentiation, erythropoiesis, and platelet biology [11–13]. These findings raise the possibility that hyperandrogenism may contribute directly to hematologic alterations observed in PCOS, independent of metabolic dysfunction.

Complete blood count (CBC)–derived indices—including the neutrophil-to-lymphocyte ratio (NLR), red cell distribution width (RDW), and platelet-to-lymphocyte ratio (PLR)—are inexpensive and widely available markers reflecting systemic inflammatory and hematologic states. Prior studies have reported inconsistent associations between these indices and PCOS [14–17], frequently without comprehensive adjustment for insulin resistance or detailed characterization of androgenic parameters. Moreover, few investigations have examined androgen-related associations with CBC-derived indices in large, relatively homogeneous cohorts of young women, in whom long-standing metabolic comorbidities may be less pronounced.

Clarifying the relative contributions of androgen excess and insulin resistance to hematologic alterations in PCOS may improve understanding of early disease mechanisms and cardiometabolic risk stratification. Therefore, we evaluated associations between androgen-related biomarkers and CBC-derived indices in a large cohort of young women with PCOS and examined whether these associations persisted after adjustment for insulin resistance.

## MATERIALS AND METHODS

### Study Population

This cross-sectional study included 1,300 consecutive women aged 16–25 years diagnosed with polycystic ovary syndrome between 2018 and 2025 at a single-center Gynecological Endocrinology Clinic in Katowice, Poland. PCOS was diagnosed according to the Rotterdam criteria, requiring at least two of the following after exclusion of related endocrine disorders: oligo- or anovulation, clinical and/or biochemical hyperandrogenism, and polycystic ovarian morphology on ultrasound.

Available-case (AC) analyses included all participants with non-missing data for a given exposure–outcome pair. Complete-case (CC) analyses were restricted to women with non-missing values for androgenic markers, homeostasis model assessment of insulin resistance (HOMA-IR), and age. Multivariable regression models adjusting for log-transformed HOMA-IR included 885–888 participants, depending on outcome-specific missingness.

### Laboratory Measurements

Fasting venous blood samples were collected in the morning after an overnight fast. Serum total testosterone and dehydroepiandrosterone sulfate (DHEAS) were measured using standardized immunoassays in a certified clinical laboratory throughout the study period, according to routine quality-control procedures.

The free androgen index (FAI) was calculated as:

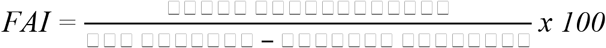

Insulin resistance was estimated using HOMA-IR, calculated from fasting glucose and insulin concentrations. Due to right-skewed distribution, HOMA-IR was log-transformed prior to inclusion in regression models.

CBC parameters were measured using automated hematology analyzers under routine laboratory quality assurance protocols. NLR and PLR were calculated from absolute cell counts. RDW was analyzed as reported by the laboratory.

### Statistical Analysis

Continuous variables are presented as medians with interquartile ranges (IQR) due to non-normal distribution. Associations between androgenic biomarkers and CBC-derived indices were first evaluated using Spearman rank correlation coefficients.

To control for multiple testing, P values were adjusted using the Benjamini–Hochberg false discovery rate (FDR) procedure within each family of related tests.

Multivariable linear regression models were fitted separately for each androgenic biomarker (FAI, total testosterone, and DHEAS) with CBC-derived indices (NLR, RDW, and PLR) as outcomes. Models were adjusted for age and log-transformed HOMA-IR.

To account for potential heteroskedasticity, heteroskedasticity-consistent (HC3) robust standard errors were used. Model diagnostics included evaluation of influential observations using Cook’s distance. Statistical significance was defined as FDR-adjusted q < 0.05.

All analyses were performed using Python (version 3.12) with standard scientific libraries (NumPy, pandas, SciPy, and statsmodels).

### Ethics Approval

The study protocol was approved by the institutional ethics committee in Poland (approval number: 254/2021). The study was conducted in accordance with the Declaration of Helsinki. Written informed consent was obtained from all participants. For participants younger than 18 years, written informed consent was obtained from a parent or legal guardian.

## RESULTS

### Study Population

The analytic cohort comprised 1,300 women with PCOS (median age 21 years [IQR 19–22]). AC analyses for primary CBC-derived indices included approximately 900–910 participants, depending on variable-specific missingness. CC analyses, restricted to participants with non-missing androgenic markers, HOMA-IR, and age, included approximately 905 women. Multivariable regression models adjusted for log-transformed HOMA-IR included 885–888 participants, depending on outcome.

Baseline characteristics were comparable between complete-case participants and those excluded due to missingness (Supplementary Table 1).

**Table 1.**
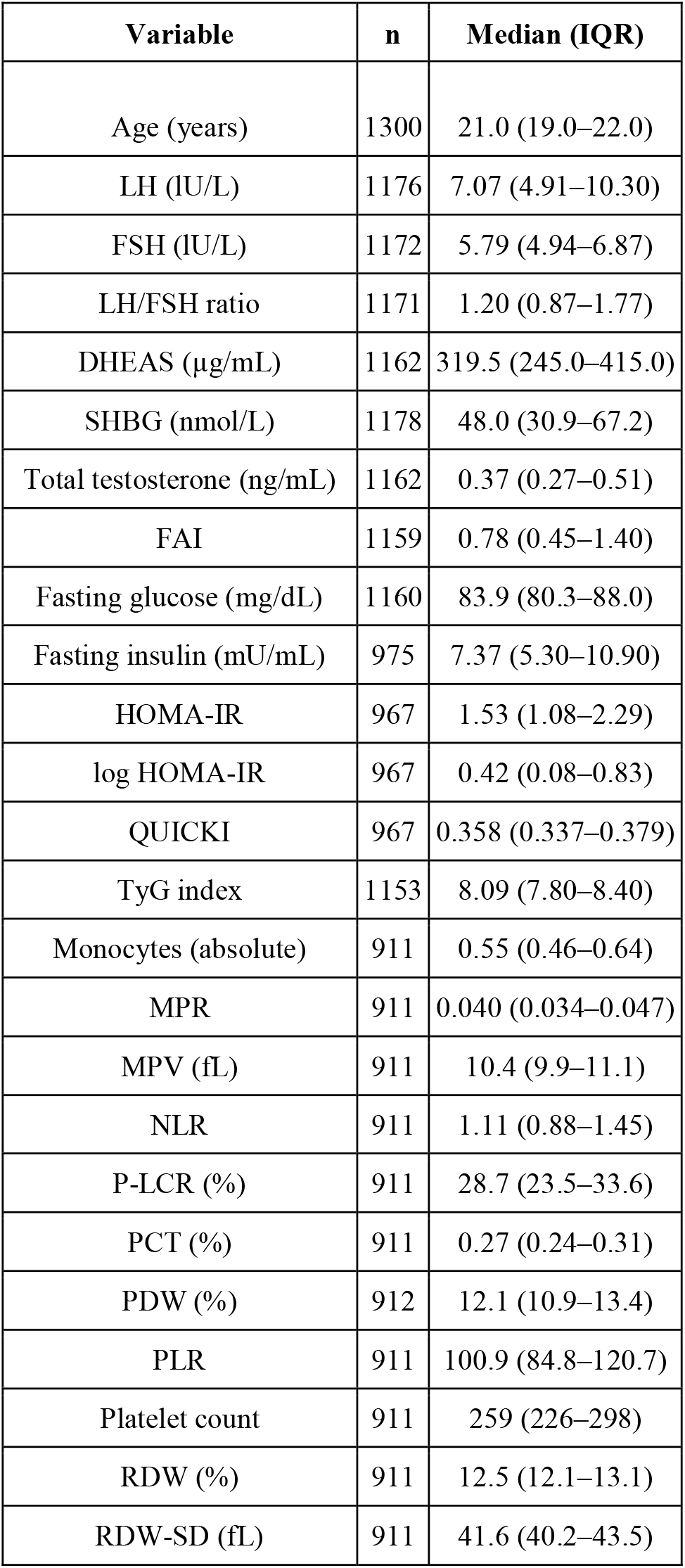
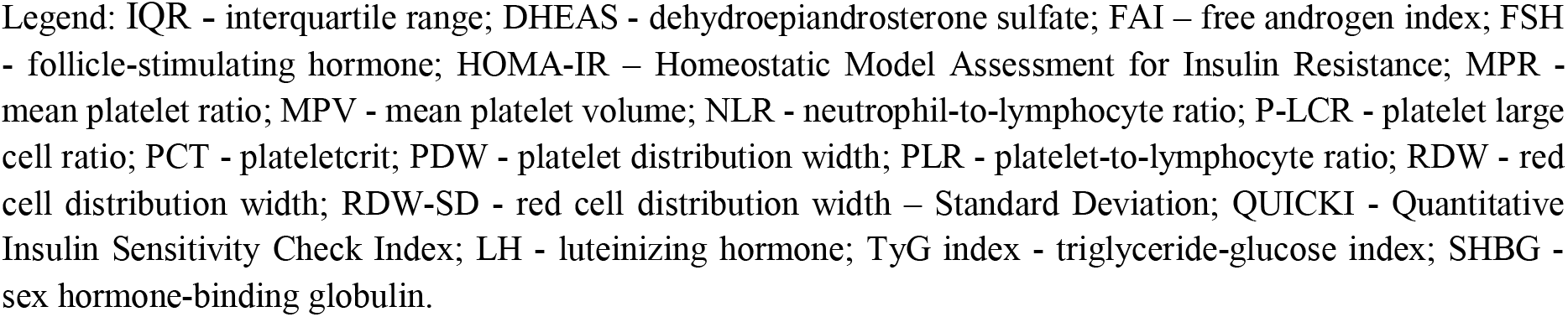
Baseline Characteristics of the Study Population (Available-Case Cohort)

| Variable | n | Median (IQR) |
| --- | --- | --- |
| Age (years) | 1300 | 21.0 (19.0–22.0) |
| LH (IU/L) | 1176 | 7.07 (4.91–10.30) |
| FSH (IU/L) | 1172 | 5.79 (4.94–6.87) |
| LH/FSH ratio | 1171 | 1.20 (0.87–1.77) |
| DHEAS (µg/mL) | 1162 | 319.5 (245.0–415.0) |
| SHBG (nmol/L) | 1178 | 48.0 (30.9–67.2) |
| Total testosterone (ng/mL) | 1162 | 0.37 (0.27–0.51) |
| FAI | 1159 | 0.78 (0.45–1.40) |
| Fasting glucose (mg/dL) | 1160 | 83.9 (80.3–88.0) |
| Fasting insulin (mU/mL) | 975 | 7.37 (5.30–10.90) |
| HOMA-IR | 967 | 1.53 (1.08–2.29) |
| log HOMA-IR | 967 | 0.42 (0.08–0.83) |
| QUICKI | 967 | 0.358 (0.337–0.379) |
| TyG index | 1153 | 8.09 (7.80–8.40) |
| Monocytes (absolute) | 911 | 0.55 (0.46–0.64) |
| MPR | 911 | 0.040 (0.034–0.047) |
| MPV (fL) | 911 | 10.4 (9.9–11.1) |
| NLR | 911 | 1.11 (0.88–1.45) |
| P-LCR (%) | 911 | 28.7 (23.5–33.6) |
| PCT (%) | 911 | 0.27 (0.24–0.31) |
| PDW (%) | 912 | 12.1 (10.9–13.4) |
| PLR | 911 | 100.9 (84.8–120.7) |
| Platelet count | 911 | 259 (226–298) |
| RDW (%) | 911 | 12.5 (12.1–13.1) |
| RDW-SD (fL) | 911 | 41.6 (40.2–43.5) |

Legend: IQR - interquartile range; DHEAS - dehydroepiandrosterone sulfate; FAI – free androgen index; FSH - follicle-stimulating hormone; HOMA-IR – Homeostatic Model Assessment for Insulin Resistance; MPR - mean platelet ratio; MPV - mean platelet volume; NLR - neutrophil-to-lymphocyte ratio; P-LCR - platelet large cell ratio; PCT - plateletcrit; PDW - platelet distribution width; PLR - platelet-to-lymphocyte ratio; RDW - red cell distribution width; RDW-SD - red cell distribution width – Standard Deviation; QUICKI - Quantitative Insulin Sensitivity Check Index; LH - luteinizing hormone; TyG index - triglyceride-glucose index; SHBG - sex hormone-binding globulin.

### Correlation Analyses

#### Inflammatory indices

In available-case analyses, FAI demonstrated a positive association with NLR (Spearman ρ = 0.201; FDR-adjusted q < 0.001). This association was consistent in complete-case analyses.

DHEAS showed a modest positive association with NLR in complete-case analyses (FDR-adjusted q = 0.040).

No other correlations between androgenic biomarkers and inflammatory indices (RDW, PLR) remained statistically significant after FDR correction.

**Table 2.** Spearman Correlations Between Androgenic Markers and CBC-Derived Indices (Available-Case and Complete-Case Analyses)

| Androgenic marker | CBC-derived index | n (AC) | Spearman $\rho$ (AC) | P value (AC) | FDR q (AC) | n (CC) | Spearman $\rho$ (CC) | P value (CC) | FDR q (CC) |
| --- | --- | --- | --- | --- | --- | --- | --- | --- | --- |
| FAI | NLR | 907 | 0.201 | <0.001 | <0.001 | 905 | 0.201 | <0.001 | <0.001 |
| FAI | PLR | 907 | -0.019 | 0.565 | 0.811 | 905 | -0.022 | 0.518 | 0.777 |
| FAI | RDW (%) | 907 | 0.001 | 0.986 | 0.986 | 905 | 0.002 | 0.954 | 0.954 |
| DHEAS ($\mu\text{g/mL}$) | NLR | 906 | 0.084 | 0.012 | 0.052 | 905 | 0.087 | 0.009 | 0.040 |
| DHEAS ($\mu\text{g/mL}$) | PLR | 906 | 0.041 | 0.216 | 0.484 | 905 | 0.043 | 0.197 | 0.443 |
| DHEAS ($\mu\text{g/mL}$) | RDW (%) | 906 | -0.016 | 0.631 | 0.811 | 905 | -0.013 | 0.688 | 0.884 |
| Total testosterone (ng/mL) | NLR | 908 | 0.064 | 0.053 | 0.159 | 905 | 0.067 | 0.043 | 0.128 |
| Total testosterone (ng/mL) | PLR | 908 | -0.037 | 0.269 | 0.484 | 905 | -0.038 | 0.254 | 0.458 |
| Total testosterone (ng/mL) | RDW (%) | 908 | -0.011 | 0.750 | 0.844 | 905 | -0.006 | 0.865 | 0.954 |

Legend: FAI – free androgen index; DHEAS - dehydroepiandrosterone sulfate; NLR - neutrophil-to-lymphocyte ratio; PLR - platelet-to-lymphocyte ratio; RDW - red cell distribution width; AC - available-case; CC - complete-case; FDR - false discovery rate.

#### Platelet morphology indices

No reproducible FDR-significant associations were observed between androgenic markers and platelet size indices (MPV, PDW, P-LCR) in either AC or CC analyses (Supplementary Table 2).

#### Multivariable Regression Analyses

Multivariable linear regression models were fitted separately for each androgenic biomarker and each CBC-derived outcome, adjusted for age and log-transformed HOMA-IR with HC3 robust standard errors.

### RDW

Higher FAI was independently associated with lower RDW(β = −0.075; 95% CI −0.141 to −0.009; FDR-adjusted q = 0.032).

Higher DHEAS was also independently associated with lower RDW (β = −0.00056; 95% CI −0.00109 to −0.00004; q = 0.042).

Total testosterone was not significantly associated with RDW after adjustment.

### PLR

Higher FAI was independently associated with lower PLR (β = −2.37; 95% CI −4.60 to −0.14; q = 0.042).

In complete-case analyses, total testosterone was inversely associated with PLR (β = −3.91; 95% CI −7.58 to −0.24; q = 0.038).

DHEAS was not significantly associated with PLR after adjustment.

### NLR

Although FAI demonstrated a significant positive association with NLR in unadjusted analyses, this association did not remain statistically significant after adjustment for age and log(HOMA-IR) (Table 3).

Similarly, no androgenic marker was independently associated with NLR in fully adjusted models after false discovery rate (FDR) correction. Full regression outputs for all crude and adjusted models are provided in Supplementary Table 3.

**Table 3.**
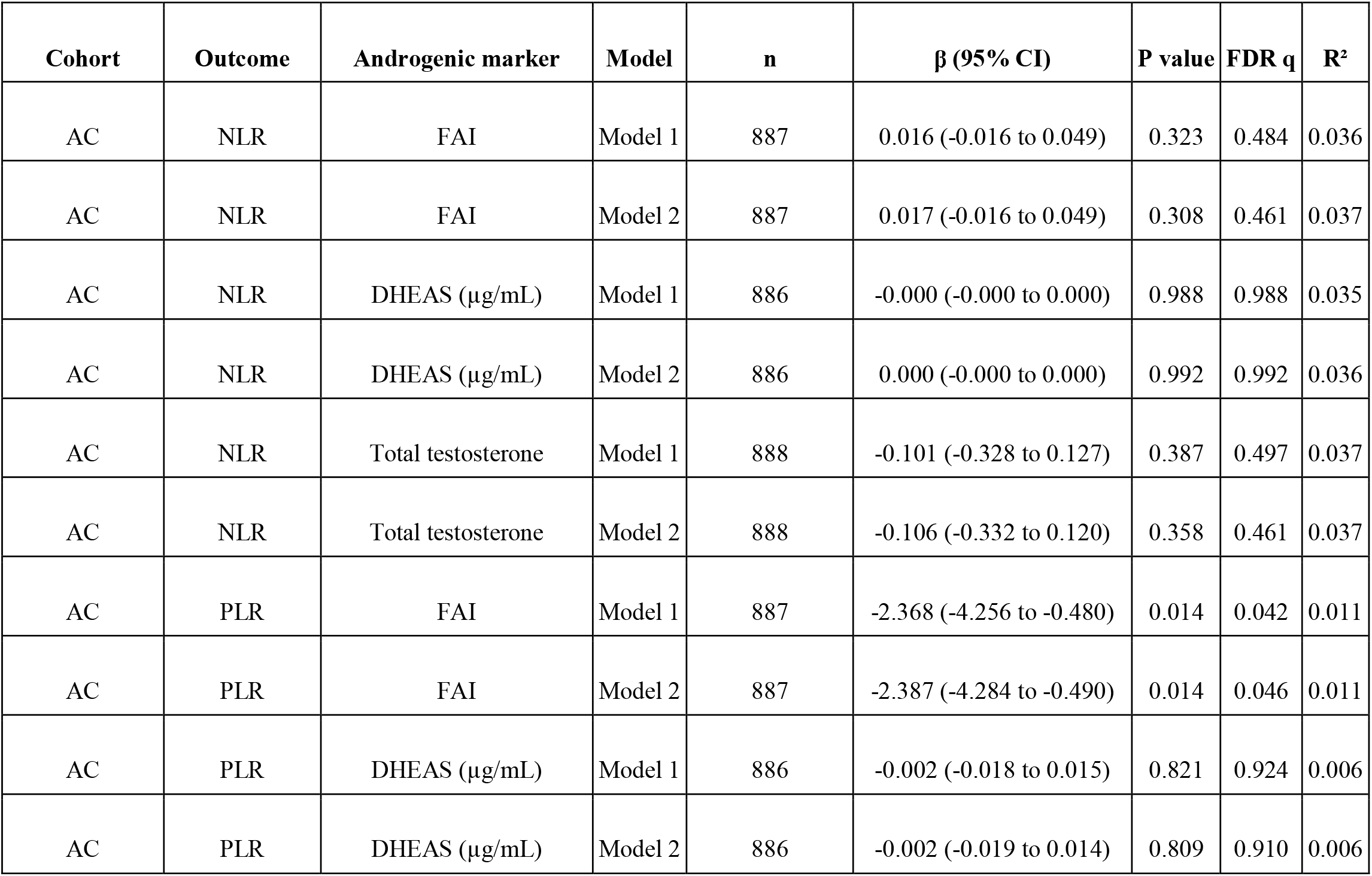
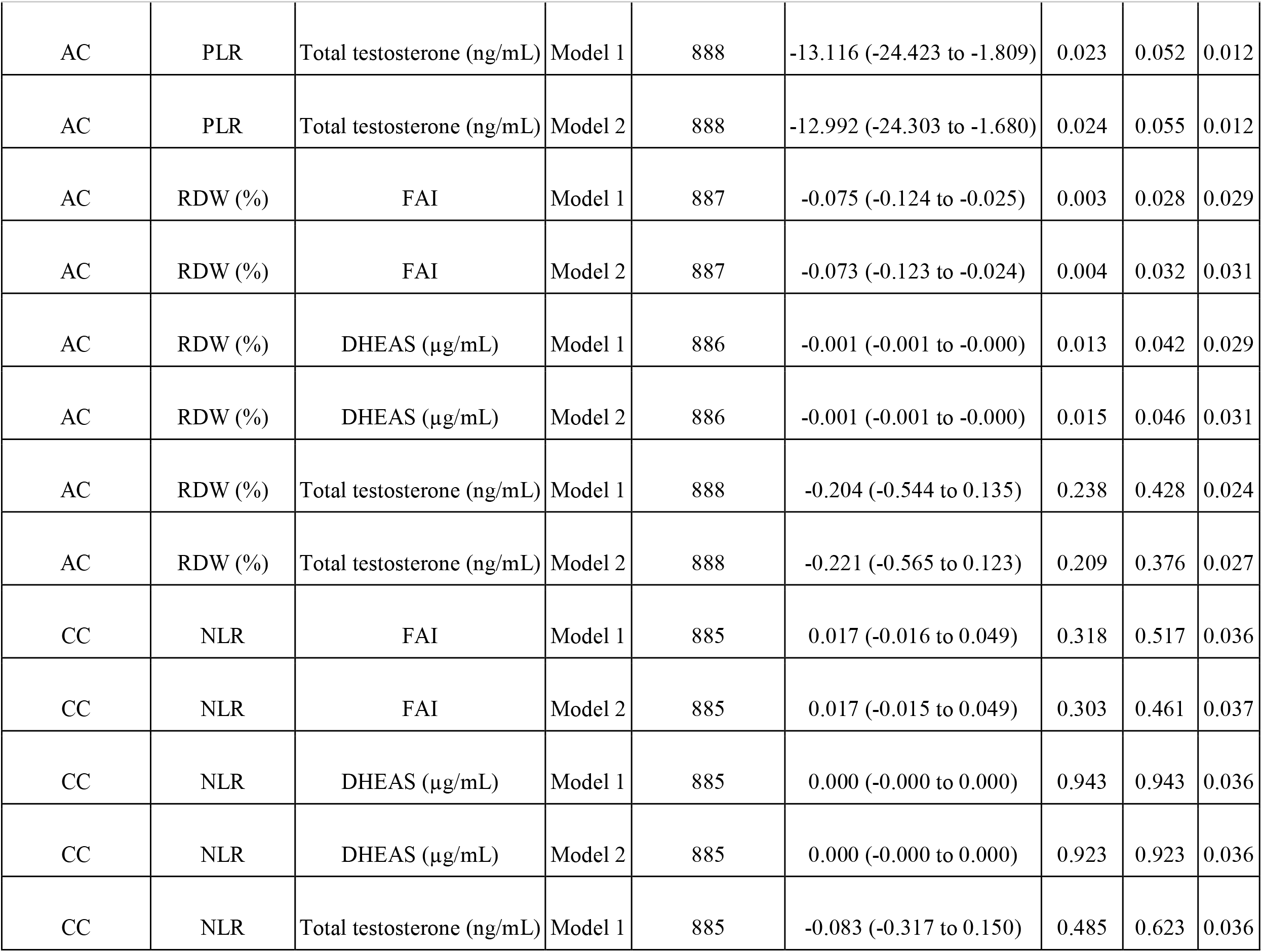
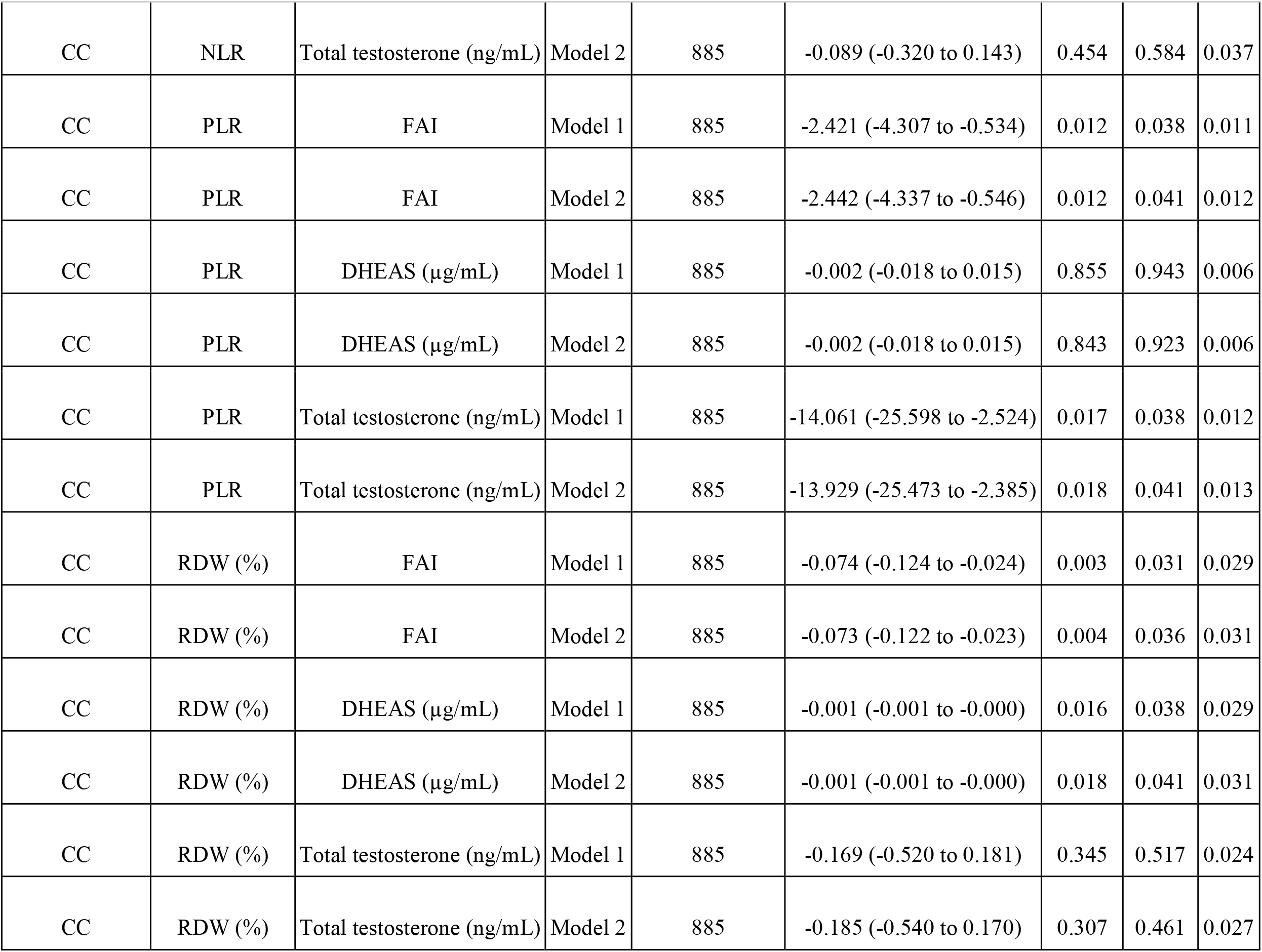
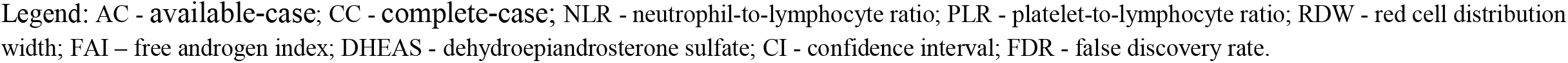
Multivariable Linear Regression Models of Androgenic Markers and CBC-Derived Indices.

| Cohort | Outcome | Androgenic marker | Model | n | $\beta$ (95% CI) | P value | FDR q | R<sup>2</sup> |
| --- | --- | --- | --- | --- | --- | --- | --- | --- |
| AC | NLR | FAI | Model 1 | 887 | 0.016 (-0.016 to 0.049) | 0.323 | 0.484 | 0.036 |
| AC | NLR | FAI | Model 2 | 887 | 0.017 (-0.016 to 0.049) | 0.308 | 0.461 | 0.037 |
| AC | NLR | DHEAS ($\mu\text{g/mL}$) | Model 1 | 886 | -0.000 (-0.000 to 0.000) | 0.988 | 0.988 | 0.035 |
| AC | NLR | DHEAS ($\mu\text{g/mL}$) | Model 2 | 886 | 0.000 (-0.000 to 0.000) | 0.992 | 0.992 | 0.036 |
| AC | NLR | Total testosterone | Model 1 | 888 | -0.101 (-0.328 to 0.127) | 0.387 | 0.497 | 0.037 |
| AC | NLR | Total testosterone | Model 2 | 888 | -0.106 (-0.332 to 0.120) | 0.358 | 0.461 | 0.037 |
| AC | PLR | FAI | Model 1 | 887 | -2.368 (-4.256 to -0.480) | 0.014 | 0.042 | 0.011 |
| AC | PLR | FAI | Model 2 | 887 | -2.387 (-4.284 to -0.490) | 0.014 | 0.046 | 0.011 |
| AC | PLR | DHEAS ($\mu\text{g/mL}$) | Model 1 | 886 | -0.002 (-0.018 to 0.015) | 0.821 | 0.924 | 0.006 |
| AC | PLR | DHEAS ($\mu\text{g/mL}$) | Model 2 | 886 | -0.002 (-0.019 to 0.014) | 0.809 | 0.910 | 0.006 |
| AC | PLR | Total testosterone (ng/mL) | Model 1 | 888 | -13.116 (-24.423 to -1.809) | 0.023 | 0.052 | 0.012 |
| AC | PLR | Total testosterone (ng/mL) | Model 2 | 888 | -12.992 (-24.303 to -1.680) | 0.024 | 0.055 | 0.012 |
| AC | RDW (%) | FAI | Model 1 | 887 | -0.075 (-0.124 to -0.025) | 0.003 | 0.028 | 0.029 |
| AC | RDW (%) | FAI | Model 2 | 887 | -0.073 (-0.123 to -0.024) | 0.004 | 0.032 | 0.031 |
| AC | RDW (%) | DHEAS (µg/mL) | Model 1 | 886 | -0.001 (-0.001 to -0.000) | 0.013 | 0.042 | 0.029 |
| AC | RDW (%) | DHEAS (µg/mL) | Model 2 | 886 | -0.001 (-0.001 to -0.000) | 0.015 | 0.046 | 0.031 |
| AC | RDW (%) | Total testosterone (ng/mL) | Model 1 | 888 | -0.204 (-0.544 to 0.135) | 0.238 | 0.428 | 0.024 |
| AC | RDW (%) | Total testosterone (ng/mL) | Model 2 | 888 | -0.221 (-0.565 to 0.123) | 0.209 | 0.376 | 0.027 |
| CC | NLR | FAI | Model 1 | 885 | 0.017 (-0.016 to 0.049) | 0.318 | 0.517 | 0.036 |
| CC | NLR | FAI | Model 2 | 885 | 0.017 (-0.015 to 0.049) | 0.303 | 0.461 | 0.037 |
| CC | NLR | DHEAS (µg/mL) | Model 1 | 885 | 0.000 (-0.000 to 0.000) | 0.943 | 0.943 | 0.036 |
| CC | NLR | DHEAS (µg/mL) | Model 2 | 885 | 0.000 (-0.000 to 0.000) | 0.923 | 0.923 | 0.036 |
| CC | NLR | Total testosterone (ng/mL) | Model 1 | 885 | -0.083 (-0.317 to 0.150) | 0.485 | 0.623 | 0.036 |
| CC | NLR | Total testosterone (ng/mL) | Model 2 | 885 | -0.089 (-0.320 to 0.143) | 0.454 | 0.584 | 0.037 |
| CC | PLR | FAI | Model 1 | 885 | -2.421 (-4.307 to -0.534) | 0.012 | 0.038 | 0.011 |
| CC | PLR | FAI | Model 2 | 885 | -2.442 (-4.337 to -0.546) | 0.012 | 0.041 | 0.012 |
| CC | PLR | DHEAS (µg/mL) | Model 1 | 885 | -0.002 (-0.018 to 0.015) | 0.855 | 0.943 | 0.006 |
| CC | PLR | DHEAS (µg/mL) | Model 2 | 885 | -0.002 (-0.018 to 0.015) | 0.843 | 0.923 | 0.006 |
| CC | PLR | Total testosterone (ng/mL) | Model 1 | 885 | -14.061 (-25.598 to -2.524) | 0.017 | 0.038 | 0.012 |
| CC | PLR | Total testosterone (ng/mL) | Model 2 | 885 | -13.929 (-25.473 to -2.385) | 0.018 | 0.041 | 0.013 |
| CC | RDW (%) | FAI | Model 1 | 885 | -0.074 (-0.124 to -0.024) | 0.003 | 0.031 | 0.029 |
| CC | RDW (%) | FAI | Model 2 | 885 | -0.073 (-0.122 to -0.023) | 0.004 | 0.036 | 0.031 |
| CC | RDW (%) | DHEAS (µg/mL) | Model 1 | 885 | -0.001 (-0.001 to -0.000) | 0.016 | 0.038 | 0.029 |
| CC | RDW (%) | DHEAS (µg/mL) | Model 2 | 885 | -0.001 (-0.001 to -0.000) | 0.018 | 0.041 | 0.031 |
| CC | RDW (%) | Total testosterone (ng/mL) | Model 1 | 885 | -0.169 (-0.520 to 0.181) | 0.345 | 0.517 | 0.024 |
| CC | RDW (%) | Total testosterone (ng/mL) | Model 2 | 885 | -0.185 (-0.540 to 0.170) | 0.307 | 0.461 | 0.027 |

Legend: AC - available-case; CC - complete-case; NLR - neutrophil-to-lymphocyte ratio; PLR - platelet-to-lymphocyte ratio; RDW - red cell distribution width; FAI – free androgen index; DHEAS - dehydroepiandrosterone sulfate; CI - confidence interval; FDR - false discovery rate.

### Model Diagnostics

Use of HC3 robust standard errors did not materially alter statistical inference. Evaluation of influential observations using Cook’s distance did not identify observations materially affecting model estimates (Supplementary Table 4).

## DISCUSSION

In this large cohort of young women with PCOS, we systematically evaluated the relationships between androgen-related biomarkers and CBC-derived inflammatory and platelet indices while carefully accounting for insulin resistance. Three principal observations emerge.

First, FAI demonstrated a positive correlation with NLR in unadjusted analyses. However, this association did not persist after adjustment for log(HOMA-IR) and age, and no androgenic marker remained independently associated with NLR after false discovery rate (FDR) correction. Prior studies examining NLR in PCOS have yielded heterogeneous results, with several reports indicating elevated NLR in PCOS populations but attributing this primarily to obesity, insulin resistance, or metabolic syndrome components rather than androgen excess per se [18–20]. Our findings are consistent with this interpretation and suggest that the apparent relationship between androgenicity and NLR may reflect overlapping metabolic pathways rather than a direct androgen-specific inflammatory effect.

Second, multivariable models identified inverse associations between androgenic markers and selected hematologic indices. Higher FAI was independently associated with lower PLR and RDW after adjustment for log(HOMA-IR), with similar estimates in available-case and complete-case analyses. Total testosterone demonstrated comparable inverse associations with PLR, whereas DHEAS showed modest but statistically significant inverse associations with RDW. Although effect sizes were small and some associations were attenuated after FDR correction, the reproducibility across analytic strategies supports internal consistency.

The inverse associations observed between androgenic markers and RDW are biologically plausible. RDW has been linked to systemic inflammation, cardiometabolic risk, and adverse cardiovascular outcomes across diverse populations [21,22]. Experimental data demonstrate androgen receptor expression in hematopoietic progenitor cells and erythroid lineages, and androgens are known to stimulate erythropoiesis [23,24]. Androgen-mediated modulation of erythroid maturation or red cell heterogeneity therefore represents a plausible mechanistic pathway. Whether lower RDW in the context of higher androgenicity reflects altered erythropoietic kinetics, iron handling, or immune–hematopoietic cross-talk remains to be determined.

Similarly, the inverse associations between androgenic markers and PLR may reflect androgen-related influences on platelet biology and lymphocyte dynamics. Platelet production and function are regulated by inflammatory and hormonal signaling pathways, and androgen receptor expression has been documented within hematopoietic tissues [23,25]. While PLR is frequently interpreted as an inflammatory surrogate, it may also integrate subtle shifts in thrombopoiesis and immune cell distribution. The modest magnitude of the observed effects suggests regulatory modulation rather than overt inflammatory activation.

Importantly, attenuation of androgen–NLR associations after metabolic adjustment underscores the central role of insulin resistance in shaping inflammatory phenotypes in PCOS. Insulin resistance has been consistently associated with low-grade inflammation and alterations in leukocyte indices [18,19]. Our findings extend prior work by demonstrating that, after rigorous adjustment and FDR correction, androgenic markers do not independently predict NLR in this age-homogeneous cohort.

### Clinical Implications

CBC-derived indices such as NLR, PLR, and RDW are inexpensive and routinely available in clinical practice. Although the associations observed were modest and do not support immediate clinical application, they may reflect subtle hematologic signatures associated with androgen excess in young women with PCOS. Whether such indices have prognostic value for long-term cardiometabolic outcomes remains uncertain and warrants prospective investigation [21,22].

### Strengths and Limitations

Strengths of this study include the large sample size, narrow age range minimizing confounding by age-related cardiometabolic progression, and comprehensive analytic strategy incorporating both available-case and complete-case approaches. The use of HC3 standard errors and FDR correction enhances statistical rigor and reduces the likelihood of false-positive inference.

Limitations include the cross-sectional design, precluding causal conclusions; incomplete anthropometric data limiting full adjustment for adiposity-related confounding; and missingness of selected platelet-related parameters. Additionally, the single-center design may limit generalizability. Finally, although several associations reached statistical significance, effect sizes were small and should be interpreted cautiously until externally validated.

## CONCLUSION

In this large cross-sectional cohort of young women with PCOS, androgen-related biomarkers were associated with selected CBC-derived hematologic indices. Although FAI demonstrated a positive correlation with NLR in unadjusted analyses, this association was not independent of insulin resistance after multivariable adjustment and FDR correction. In contrast, higher androgenic markers were reproducibly associated with lower RDW and PLR in adjusted models, with consistent estimates across available-case and complete-case analyses.

These findings suggest that androgen excess in PCOS may be linked to subtle modulation of erythroid and platelet-related parameters beyond the effects of insulin resistance. However, the observed effect sizes were modest, and causal inference cannot be established. Longitudinal studies are required to determine whether these hematologic alterations have clinical or prognostic significance.

## Data Availability

All data produced in the present study are available upon reasonable request to the authors.

## DATA AVAILABILITY

The datasets generated and analyzed during the current study are not publicly available due to institutional ethical regulations and data protection policies. De-identified data may be made available from the corresponding author upon reasonable request and subject to institutional approval.

The statistical analysis code used to generate the results reported in this article is publicly available at GitHub: https://github.com/npiorkowska-science/pcos-androgen-cbc-inflammation

## AUTHOR CONTRIBUTIONS

AB, KB conceptualized and designed the study.

NP performed the statistical analyses, developed the analysis code, conducted the data analyses, and drafted the initial manuscript.

GF, AB, KB contributed to the intellectual content of the study, critically revised the manuscript, and provided final editorial oversight.

All authors reviewed and approved the final version of the manuscript.

## ACKNOWLEDGMENTS

The authors thank the clinical and laboratory staff involved in data collection and patient care.

## SUPPLEMENTARY MATERIALS

### SUPPLEMENTARY TABLE

Supplementary Table 1. Comparison of Included vs Excluded Participants

Supplementary Table 2. Correlations Between Androgenic Markers and Platelet Morphology Indices

Supplementary Table 3. Full Regression Model Outputs

Supplementary Table 4. Cook’s Distance Diagnostics

## REFERENCES

1. Rotterdam ESHRE/ASRM-Sponsored PCOS Consensus Workshop Group. Revised 2003 consensus on diagnostic criteria and long-term health risks related to polycystic ovary syndrome (PCOS). Fertil Steril. 2004;81(1):19–25. doi:10.1016/j.fertnstert.2003.10.004.

2. Teede HJ, Misso ML, Costello MF, et al. Recommendations from the international evidence-based guideline for the assessment and management of polycystic ovary syndrome. Hum Reprod. 2018;33(9):1602–1618. doi:10.1093/humrep/dey256.

3. Wekker V, van Dammen L, Koning A, et al. Long-term cardiometabolic disease risk in women with PCOS: a systematic review and meta-analysis. Hum Reprod Update. 2020;26(6):942–960. doi:10.1093/humupd/dmaa029.

4. Azziz R, Carmina E, Chen Z, et al. Polycystic ovary syndrome. Nat Rev Dis Primers. 2016;2:16057. doi:10.1038/nrdp.2016.57.

5. Escobar-Morreale HF, Luque-Ramírez M, González F. Circulating inflammatory markers in polycystic ovary syndrome: a systematic review and metaanalysis. Fertil Steril. 2011;95(3):1048-1058.e1-2. doi:10.1016/j.fertnstert.2010.11.036.

6. Aboeldalyl S, James C, Seyam E, Ibrahim EM, Shawki HE, Amer S. The role of chronic inflammation in polycystic ovarian syndrome: a systematic review and meta-analysis. Int J Mol Sci. 2021;22(5):2734. doi:10.3390/ijms22052734.

7. González F. Inflammation in polycystic ovary syndrome: underpinning of insulin resistance and ovarian dysfunction. Steroids. 2012;77(4):300–305. doi:10.1016/j.steroids.2011.12.003.

8. González F, Rote NS, Minium J, Kirwan JP. Reactive oxygen species-induced oxidative stress in PCOS: implications for insulin resistance. J Clin Endocrinol Metab. 2006;91(1):336–340. doi:10.1210/jc.2005-1465.

9. Ben-Batalla I, Vargas-Delgado ME, von Amsberg G, Janning M, Loges S. Influence of androgens on immunity to self and foreign: effects on immunity and cancer. Front Immunol. 2020;11:1184. doi:10.3389/fimmu.2020.01184.

10. Mantalaris A, Panoskaltsis N, Sakai Y, et al. Localization of androgen receptor expression in human bone marrow. J Pathol. 2001;193(3):361–366. doi:10.1002/1096-9896(2000)

11. Warren AM, Grossmann M. Haematological actions of androgens. Best Pract Res Clin Endocrinol Metab. 2022;36(5):101653. doi:10.1016/j.beem.2022.101653.

12. Bachman E, Feng R, Travison T, et al. Testosterone suppresses hepcidin in men: a potential mechanism for testosterone-induced erythrocytosis. J Clin Endocrinol Metab. 2010;95(10):4743–4747. doi:10.1210/jc.2010-0864.

13. Bray PF. Androgen receptor expression in megakaryocytes and platelets. Blood. 2000;95(7):2289–2296.

14. Li L, Yu J, Zhou Z. Association between neutrophil-to-lymphocyte ratio and polycystic ovary syndrome: a PRISMA-compliant systematic review and meta-analysis. Medicine (Baltimore). 2022;101(36):e30512. doi:10.1097/MD.0000000000030512.

15. Pergialiotis V, Trakakis E, Parthenis C, et al. Correlation of platelet to lymphocyte and neutrophil to lymphocyte ratio with hormonal and metabolic parameters in women with PCOS. Horm Mol Biol Clin Investig. 2018;34(3):20170073. doi:10.1515/hmbci-2017-0073.

16. Yilmaz MA, Duran C, Basaran M. The mean platelet volume and neutrophil-to-lymphocyte ratio in obese and lean patients with polycystic ovary syndrome. J Endocrinol Invest. 2016;39(1):45–53. doi:10.1007/s40618-015-0335-2.

17. Alhabardi NAA, Al-Wutayd O, Eltayieb KM, et al. Peripheral hematological parameters in women with polycystic ovary syndrome. J Int Med Res. 2020;48(9):300060520952282. doi:10.1177/0300060520952282.

18. Keskin Kurt R, Okyay AG, Hakverdi AU, Gungoren A, Dolapcioglu KS, Karateke A. The effect of obesity on inflammatory markers in patients with polycystic ovary syndrome: a BMI-matched case-control study. Arch Gynecol Obstet. 2014;290(2):315–319. doi:10.1007/s00404-014-3199-3.

19. Kahal H, Aburima A, Ungvari T, et al. Polycystic ovary syndrome has no independent effect on vascular, inflammatory or thrombotic markers when matched for obesity. Clin Endocrinol (Oxf). 2013;79(2):252–258. doi:10.1111/cen.12137.

20. Liu W, Yin Y, Chen X, et al. Neutrophil-to-lymphocyte ratio and platelet-to-lymphocyte ratio in patients with polycystic ovary syndrome: a retrospective study. Medicine (Baltimore). 2022;101(25):e29678. doi:10.1097/MD.0000000000029678.

21. Huang YL, Hu ZD, Liu SJ, et al. Prognostic value of red blood cell distribution width for patients with heart failure: a systematic review and meta-analysis of cohort studies. PLoS One. 2014;9(8):e104861. doi:10.1371/journal.pone.0104861.

22. Bao D, Luo G, Kan F, et al. Prognostic value of red cell distribution width in patients undergoing percutaneous coronary intervention: a meta-analysis. BMJ Open. 2020;10(9):e033378. doi:10.1136/bmjopen-2019-033378.

23. Huang CK, Lai KP, Luo J, et al. Androgen receptor differential roles in stem/progenitor cells including hematopoietic lineages. Stem Cells. 2014;32(9):2299–2308. doi:10.1002/stem.1722.

24. Kim SW, Hwang JH, Cheon JM, et al. Direct and indirect effects of androgens on survival of hematopoietic progenitor cells in vitro. J Korean Med Sci. 2005;20(3):409–416. doi:10.3346/jkms.2005.20.3.409.

25. Khetawat G, Faraday N, Nealen ML, et al. Human megakaryocytes and platelets contain the estrogen receptor β and androgen receptor (AR): testosterone regulates AR expression. Blood. 2000;95(7):2289–2296. doi:10.1182/blood.V95.7.2289.

