## Supplementary Table S3. Missingness and QC. for "Androgen-Related Associations With CBC-Derived Inflammatory Indices in Young Women With Polycystic Ovary Syndrome" (from another version of this preprint: version 1, 2025-12-26)

| **Variable** | **Total n** | **Non-missing n** | **Missing n** | **Missing %** |
| --- | --- | --- | --- | --- |
| CRP | 1315 | 19 | 1296 | 98.5551 |
| log(CRP) | 1315 | 19 | 1296 | 98.5551 |
| MPV | 1315 | 46 | 1269 | 96.5019 |
| P-LCR | 1315 | 46 | 1269 | 96.5019 |
| PCT | 1315 | 46 | 1269 | 96.5019 |
| PDW | 1315 | 46 | 1269 | 96.5019 |
| PLT | 1315 | 46 | 1269 | 96.5019 |
| RDW | 1315 | 46 | 1269 | 96.5019 |
| RDW-SD | 1315 | 46 | 1269 | 96.5019 |
| WBC | 1315 | 46 | 1269 | 96.5019 |
| EOS % | 1315 | 918 | 397 | 30.1901 |
| IG % | 1315 | 918 | 397 | 30.1901 |
| LYMPH % | 1315 | 918 | 397 | 30.1901 |
| MONO % | 1315 | 918 | 397 | 30.1901 |
| NEUT % | 1315 | 918 | 397 | 30.1901 |
| NLR | 1315 | 918 | 397 | 30.1901 |
| HOMA-IR | 1315 | 1062 | 253 | 19.2395 |
| log(HOMA-IR) | 1315 | 1062 | 253 | 19.2395 |
| Age | 1315 | 1315 | 0 | 0 |
