## Supplementary Table S2. Secondary CBC analyses and missingness. for "Androgen-Related Associations With CBC-Derived Inflammatory Indices in Young Women With Polycystic Ovary Syndrome" (from another version of this preprint: version 1, 2025-12-26)

| **y** | **x** | **n** | **r** | **p** | **q** |
| --- | --- | --- | --- | --- | --- |
| NEUT % | log(HOMA-IR) | 900 | 0.2474 | 5.1133e14 | 6.1360e14 |
| LYMPH % | log(HOMA-IR) | 900 | -0.2079 | 3.0388e10 | 1.8233e9 |
| NLR | log(HOMA-IR) | 900 | 0.1938 | 4.5773e9 | 1.8309e8 |
| NeUT % | HOMA-IR | 900 | 0.1906 | 8.2787e9 | 2.4836e8 |
| IG % | log(HOMA-IR) | 900 | 0.1594 | 1.5360e6 | 3.1966e6 |
| NLR | HOMA-IR | 900 | 0.1591 | 1.5983e6 | 3.1966e6 |
| LYMPH % | HOMA-IR | 900 | -0.1520 | 4.6651e6 | 7.9974e6 |
| IG % | HOMA-IR | 900 | 0.1253 | 2.0000e4 | 2.0000e4 |
| MONO % | log(HOMA-IR) | 900 | -0.1001 | 2.6000e3 | 3.5000e3 |
| MONO % | HOMA-IR | 900 | -0.0985 | 3.1000e3 | 3.7000e3 |
| EOS % | log(HOMA-IR) | 900 | -0.0931 | 5.2000e3 | 5.6000e3 |
| EOS % | HOMA-IR | 900 | -0.0817 | 1.4100e2 | 1.4100e2 |
