## Supplementary Table S1. Baseline characteristics of the study population and data availability for "Androgen-Related Associations With CBC-Derived Inflammatory Indices in Young Women With Polycystic Ovary Syndrome" (from another version of this preprint: version 1, 2025-12-26)

| **Blood Test** | **Total** | **Nonmissing** | **Missing** | **Missing n %** | **Mean** | **Standard deviation** | **Median** | **IQR** | **Min** | **Max** |
| --- | --- | --- | --- | --- | --- | --- | --- | --- | --- | --- |
| age | 1315 | 1315 | 0 | 0 | 20.90 | 1.95 | 21 | 3 | 16 | 25 |
| CRP | 1315 | 19 | 1296 | 98.55 | 4.72 | 13.06 | 0.30 | 3.55 | 0.10 | 58 |
| EOS % | 1315 | 918 | 397 | 30.19 | 2.92 | 1.82 | 2.50 | 1.90 | 0 | 15.90 |
| HOMA-IR | 1315 | 1062 | 253 | 19.23 | 1.89 | 1.46 | 1.51 | 1.16 | 0.15 | 20.90 |
| IG % | 1315 | 918 | 397 | 30.19 | 0.24 | 0.14 | 0.20 | 0.10 | 0 | 1 |
| log(CRP) | 1315 | 19 | 1296 | 98.55 | 0.94 | 1.01 | 0.26 | 1.31 | 0.09 | 4.07 |
| log(HOMA-IR) | 1315 | 1062 | 253 | 19.24 | 0.99 | 0.36 | 0.92 | 0.44 | 0.14 | 3.09 |
| LYMPH % | 1315 | 918 | 397 | 30.19 | 40.95 | 7.90 | 41.25 | 10.70 | 11.20 | 66.20 |
| MONO % | 1315 | 918 | 397 | 30.19 | 8.72 | 1.85 | 8.60 | 2.50 | 4.40 | 16.40 |
| MPV | 1315 | 46 | 1269 | 96.50 | 10.49 | 0.85 | 10.40 | 1.22 | 8.80 | 12.90 |
| NEUT % | 1315 | 918 | 397 | 30.19 | 46.50 | 8.24 | 46.35 | 10.97 | 22.80 | 79.20 |
| NLR | 1315 | 918 | 397 | 30.19 | 1.24 | 0.58 | 1.11 | 0.57 | 0.37 | 6.31 |
| P-LCR | 1315 | 46 | 1269 | 96.50 | 28.94 | 7.045 | 28.55 | 10.62 | 14.50 | 48.60 |
| PCT | 1315 | 46 | 1269 | 96.50 | 0.29 | 0.06 | 0.28 | 0.08 | 0.19 | 0.45 |
| PDW | 1315 | 46 | 1269 | 96.50 | 12.18 | 1.90 | 12 | 2.60 | 8.70 | 19.10 |
| PLT | 1315 | 46 | 1269 | 96.50 | 276.52 | 61.96 | 263.50 | 88.25 | 169 | 432 |
| RDW | 1315 | 46 | 1269 | 96.50 | 13.01 | 1.28 | 12.55 | 1.37 | 10.60 | 17.60 |
| RDW-SD | 1315 | 46 | 1269 | 96.50 | 41.03 | 2.90 | 40.45 | 3.85 | 36.10 | 52.10 |
| WBC | 1315 | 46 | 1269 | 96.50 | 6.77 | 1.93 | 6.39 | 2.18 | 3.77 | 13.49 |
| GLUCOSE 0’ | 1315 | 1174 | 141 | 0.89 | 84.34 | 6.07 | 83.90 | 7.77 | 51.90 | 108 |
| INSULIN 0’ | 1315 | 1069 | 246 | 0.81 | 9.02 | 6.67 | 7.30 | 5.41 | 0.20 | 98.20 |
